# Disparities in COVID-19 Reported Incidence, Knowledge, and Behavior

**DOI:** 10.1101/2020.05.15.20095927

**Authors:** Marcella Alsan, Stefanie Stantcheva, David Yang, David Cutler

## Abstract

**Background:** Data from the COVID-19 pandemic in the United States show large differences in hospitalizations and mortality across race and geography. However, there is limited data on health information, beliefs, and behaviors that might indicate different exposure to risk.

**Methods:** A sample of 5,198 respondents in the United States (80% population representative, 20% oversample of hotspot areas in New York City, Seattle, New Orleans, and Detroit) was conducted from March 29th to April 13th to measure differences in knowledge, beliefs and behavior regarding COVID-19. Linear regression was used to understand racial, geographic, political, and socioeconomic differences in COVID-19 reported incidence knowledge, and behaviors after adjusting for state-specific and survey date fixed effects.

**Results:** The largest differences in COVID-19 knowledge and behaviors are associated with race/ethnicity, gender, and age. African-Americans, men, and people <55 years old are less likely to know how the disease is spread, less likely to know symptoms of COVID-19, wash their hands less frequently, and leave the home more often. Differences by income, political orientation, and living in a hotspot area are much smaller.

**Conclusions:** There are wide gaps in COVID-19 reported incidence, knowledge regarding disease spread and symptoms, and in social distancing behavior. The findings suggest more effort is needed to increase accurate information and encourage appropriate behaviors among minority communities, men, and younger people.

## INTRODUCTION

Rates of COVID-19 differ greatly across the population. CDC data on COVID-19 hospitalization and mortality rates show a higher incidence among racial and ethnic minorities, males, the elderly, and people living in less dense areas.^1^ These data are consistent with media reports from specific areas of higher hospitalization and death rates from COVID-19 among African-Americans and lower SES individuals.^2,3,4,5,6,7^

Public health efforts to reduce COVID-19 spread will thus need to focus particularly on these traditionally disadvantaged population groups. However, such efforts have historically had more difficulty making inroads in marginalized groups. For example, rates of smoking and obesity are higher among lower socioeconomic status groups, and trust in healthcare is lower among racial minorities.^8,9,10^ Further, several polls have demonstrated a partisan divide whereby right-leaning media outlets have and individuals who consume them are skeptical about the threat posed by COVID-19 and of efforts to reduce its spread.^11,12^ This raises concerns that the COVID-19 response will be hampered by differences in knowledge, beliefs, and behaviors among racial/ethnic, socioeconomic, or groups with differing political orientations in the population.

We conducted a large-scale survey in the United States in late March and early April 2020, during the early exponential growth phase of the epidemic. The survey is national representative with an oversample of hotspot areas at the time. The survey asked questions about reported prevalence, knowledge of COVID-19, beliefs about the virus, and behaviors related to the spread of COVID-19. Results are used to determine how these measures differ by race/ethnicity, socioeconomic status, and political orientation.

## METHODOLOGY

### Data

We developed and conducted a survey on the effects of COVID-19 including 5,198 individuals. The survey was conducted in the United States from March 29 to April 13 and was carried out online by Dynata Corporation. Eighty percent of respondents were sampled from geographies according to population representation, and 20% were sampled from hotspots area, those with the highest counts of COVID-19 at the time: Seattle, New York City, New Orleans, and Detroit. Exact question wording along with summary statistics on the sample are reported in the Appendix. The survey was conducted by internet. The survey sample is very similar to the overall US population, with the exception of a higher share of individuals with a college degree in the survey than in the nation.

### Questions about COVID-19 Interactions

Two measures of direct interactions with COVID-19 were asked: whether the respondent has already contracted COVID-19, and whether the respondent personally knows someone who has contracted COVID-19.

### Questions about Knowledge and Behaviors

A number of questions were asked about knowledge of COVID-19. One question asked whether a person could infect others without being sick or showing symptoms. A separate question asked respondents to answer yes/no to five questions about COVID-19 spread: whether the virus spreads through close contact with an infected person (within about 6 feet); through respiratory droplets produced when an infected person coughs or sneezes; by touching a contaminated surface and then touching your eyes nose or mouth; though unprotected sex; and whether the virus is a hoax. Analysis focuses particularly on knowledge of fomite transmission (a binary variable for whether the respondent knows that touching a contaminated surface can lead to transmission)

Respondents were also asked to identify the top 3 symptoms of COVID-19 from a list: fever; dry eyes; skin rash; cough; difficulty breathing; swollen legs; acid reflux; stomach ache; watery eyes. From these responses, a binary variable is created for whether the respondent knows all three COVID-19 symptoms (fever, cough, and difficulty in breathing).

Behaviors to mitigate the spread of COVID-19 are measured by two variables: how many times the person washed their hands in the past 24 hours, and how many times the person left their house in the past 3 days.

#### Covariates

Reported infection, knowledge of spread and symptoms, and behaviors are related to socioeconomic and political orientation variables. Demographic variables include age (included in regression models as <30, 30-54, 55-64 and ≥65), gender (male/female), race/ethnicity (non-Hispanic white, non-Hispanic black, Hispanic), income (included in regression models as <$25,000, $25,000-$49,999, $50,000-74,999, $75,000-$99,999 and ≥$100,000), and political orientation (Democratic, Republican, Independent/No Affiliation). A dummy variable is also included for whether the individual lived in a hotspot area (New York City, Seattle, New Orleans and Detroit).

The trial was approved by the institutional review board of Harvard University and deemed exempt.

### Analysis

Linear regression analysis is used to relate measures of reported incidence, knowledge, and behaviors to the independent variables noted above. Additional controls include whether the individual has health insurance, whether the respondent reports a condition that places them at higher risk of death condition on infection (cardiovascular disease, chronic lung disease and diabetes), a measure of risk tolerance (a 0-10 measure of willingness to take risk), exact date of survey, and state of residence to account for differences in policies across geographies.^13^ Logit and probit models gave similar marginal effects, but OLS estimates are presented for ease of interpretation (see the Appendix).

## RESULTS

Table A1 compares the survey sample demographics to those in the US as a whole. The survey sample is similar to the overall U.S. population in terms of sex, age distribution, racial and employment characteristics, though the percentage of college graduates is higher than in the country as a whole.

Table 1 shows means for the measures of disease exposure, knowledge, beliefs, and behaviors. 4% of people report being infected with COVID-19, and 24% report knowing someone who has been infected with COVID-19. Knowledge about the spread of COVID-19 is very high. 83% of people believe that COVID-19 can be obtained from a contaminated surface, and 87% know all three symptoms of COVID-19 (fever, cough, difficulty in breathing). Very few people (10.9%) associate COVID-19 with sexual transmission. Only 5.1% of people believe that COVID-19 is a hoax.

**Table 1.** Survey Summary Statistics of Major Outcomes: Full Sample and by “Hotspot”

|  | Entire Sample<br>(N=5,198) | Inside Hotspot<br>(N=1,082) | Outside Hotspot<br>(N=4,116) |
| --- | --- | --- | --- |
| **Knowledge about Spread (%)** |  |  |  |
| Through Respiratory Droplets | 86% | 83% | 86% |
| Through Close Contact | 78% | 76% | 78% |
| Through a Contaminated Surface | 83% | 81% | 84% |
| Spread without Showing Symptoms | 94% | 93% | 95% |
| Through unprotected sex | 10.9% | 13.7% | 10.2% |
| The virus is a hoax | 5.1% | 8.2% | 4.3% |
| **Knowledge about Symptoms (%)** |  |  |  |
| Fever | 94% | 92% | 94% |
| Cough | 94% | 92% | 95% |
| Difficulty in Breathing | 92% | 90% | 93% |
| All Three Symptoms | 87% | 84% | 88% |
| Any other symptoms reported | 10% | 12% | 9% |
| **Reportedly COVID-19+ (%)** |  |  |  |
| Reports COVID-19 Infection (Self) | 4% | 7% | 3% |
| Reports COVID-19 Infection (Other) | 23% | 40% | 19% |
| **Subjective Likelihood** |  |  |  |
| Likelihood of getting sick from COVID-19 in next month (0-10) | 3.9<br>(2.5) | 4.1<br>(2.5) | 3.9<br>(2.5) |
| Number of community members likely to get sick from COVID-19 in next month (0-100) | 37.6<br>(30.2) | 46.7<br>(30.5) | 35.2<br>(29.7) |
| **Behaviors<sup>††</sup>** |  |  |  |
| Handwashing in Last 24 Hours | 13.2<br>(13.5) | 13.0<br>(13.0) | 13.2<br>(13.6) |
| Left Home in 3 Days | 2.5<br>(4.0) | 2.8<br>(4.6) | 2.4<br>(3.8) |
| **Frequent Hospital Use (%)** | 13% | 17% | 12% |
| **Ease of Accessing Testing (0-10)<sup>††</sup>** | 4.2 | 4.3 | 4.2 |
| (0=extremely difficult; 10=extremely easy) | (2.9) | (3.1) | (2.9) |

<sup>†</sup> N is the number eligible to answer. A small number did not answer each question.
<sup>††</sup> Mean (standard deviation)

Figure 1 shows the differential impact of demographics, socioeconomic status, geographic location, and political orientation on the probability of being COVID-19+ or knowing someone who is. African-American respondents are 3.5 percentage points (p=.001) more likely than whites to report being infected with COVID-19 as are men (3.2 percentage points, p<.001). Individuals at the top of the income distribution are 1.6 percentage points (p=.068) more likely to report infection than are those in the lowest income group. Those who affiliate with the Republican party are 2.6 percentage points (p<.001) more likely to report infection than are political independents.

**Figure 1:**
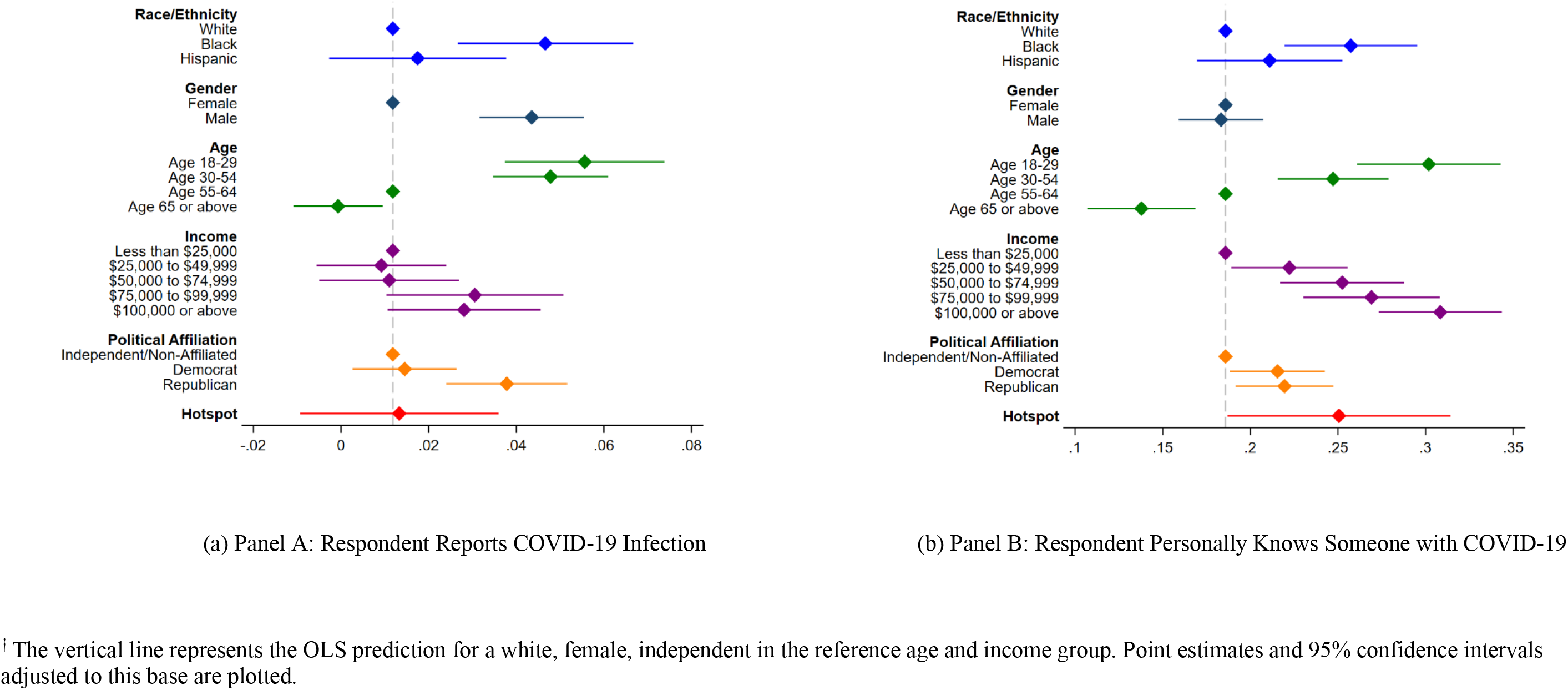
Reported COVID-19+ Infection^†^

Turning to the question on infection of acquaintances in panel (b), respondents in a “hotspot” area are more likely to know somehow who is COVID-19+ than are people living in other areas (6.5 percentage points p=.033). As above, African-Americans are far more likely to report knowing someone who is COVID-19+ than are whites (7.2 percentage points; p=.019). More young people know someone who is COVID-19+ (coefficient on age 18-29 = 11.6 percentage points; p=.021) as do higher income people (coefficient on earning $100,000 or above = 12.3 percentage points; p=.018). Despite the partisan nature of reaction to the pandemic, Democrats and Republicans are approximately equally more likely to know someone who is COVID-19+ that non-affiliated/independents.

Figure 2 shows the factors associated with higher knowledge about the spread of COVID-19. The largest gaps in knowledge are among racial/ethnic minorities, men, and young people (<55 years of age). African-Americans are 9.4 percentage points less likely to know a person can become infected by touching a contaminated surface (p=.019). Hispanic respondents are also 4.8 percentage points less likely to understand fomite spread (p=.020). Both estimates are statistically different from that of non-Hispanic white respondents. Male respondents are 5.1 percentage points (p <.001) less likely to know this information than female respondents, and the youngest age group (<30) is 10.3 percentage points (p<.001) to be aware than the reference age group (55-64). There are also differences in knowledge about COVID-19 by political affiliation and income. Richer people know more about COVID-19 than do poorer people (difference between income ≥$100,000 and income <$25,000 = 4.2 percentage points p-value = .016), whereas Republicans are less likely (difference between Republicans and non-aligned/independent = 3.3 percentage points; p-value = .015). The appendix shows additional results on asymptomatic transmission as well as transmission via respiratory droplets or contact within an infected person. The results are overall consistent with those reported in the main text.

**Figure 2:**
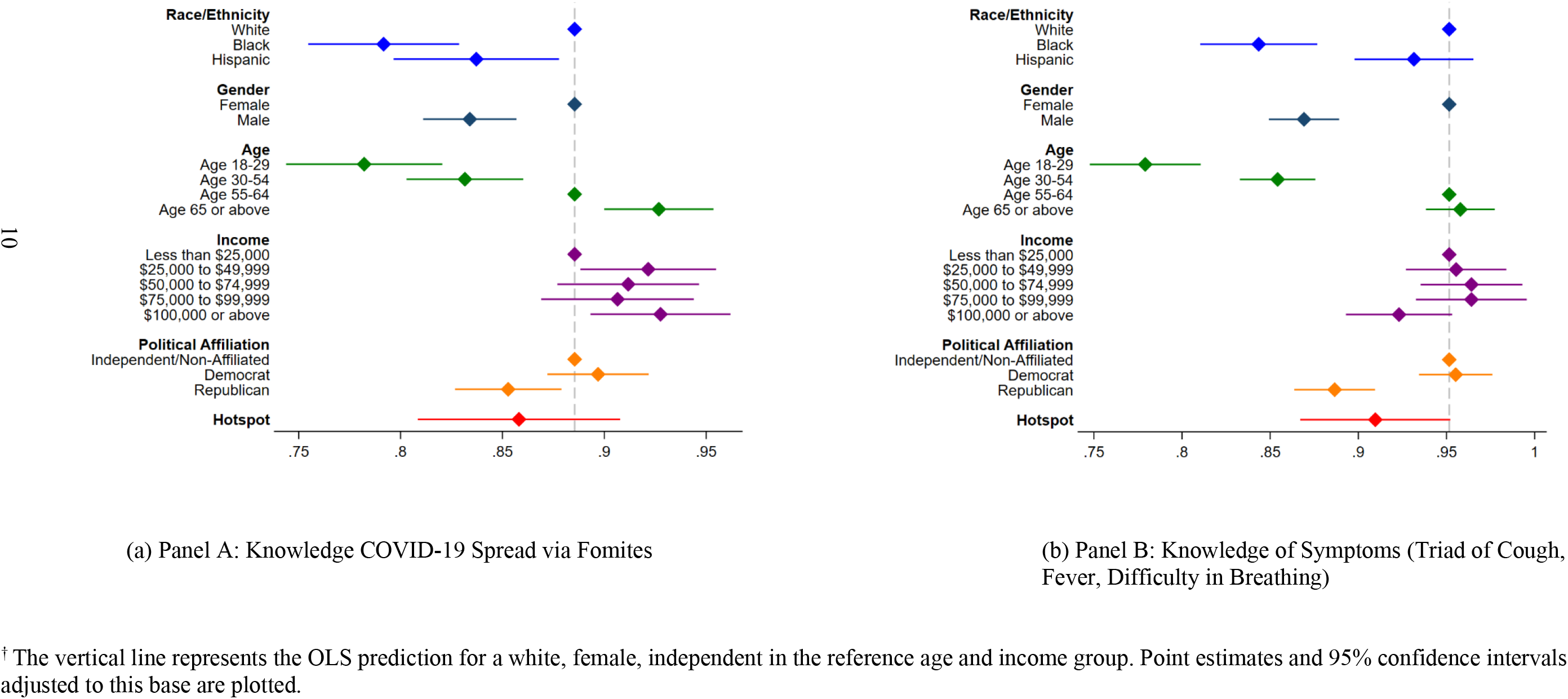
Knowledge of Spread and Symptoms^†^

Very similar effects are found with respect to knowledge of COVID-19 symptoms. African-American and Hispanic respondents are 10.8 (p <.001) and 2.0 (p = .244) percentage points less likely to know cough, fever and shortness of breath are symptoms of COVID-19 than are non-Hispanic whites. Similarly, people aged 18-29 are 17.2 percentage points (p<.001) less likely to know the symptoms of COVID-19 than are people aged 55-64.

Figure 3 shows differences in handwashing and leaving the house for the same groups. With respect to handwashing, the largest differences are between men and women and between younger and older groups. Men wash their hands 3.8 times less per 24 hours than women (p <.001) and people aged 18-29 wash their hands 4.4 times less per 24 hours than older individuals (p <.001). African-American and Hispanic respondents, on the other hand, wash their hands more frequently than white respondents in a 24 hours period (1.0 times more for the former (p = .650) and 1.8 times more for the latter (p=.020)). Men are also more likely to leave the house frequently than are women (.74 times, p<.001), as are African Americans compared to white respondents (.93 times, p <.001). Older Americans (age 65+) are less likely to leave their homes frequently (-.36 times, p = .018). Neither frequent handwashing nor staying indoors more commonly differ by income, residence in a hotspot area, or political orientation.

**Figure 3:**
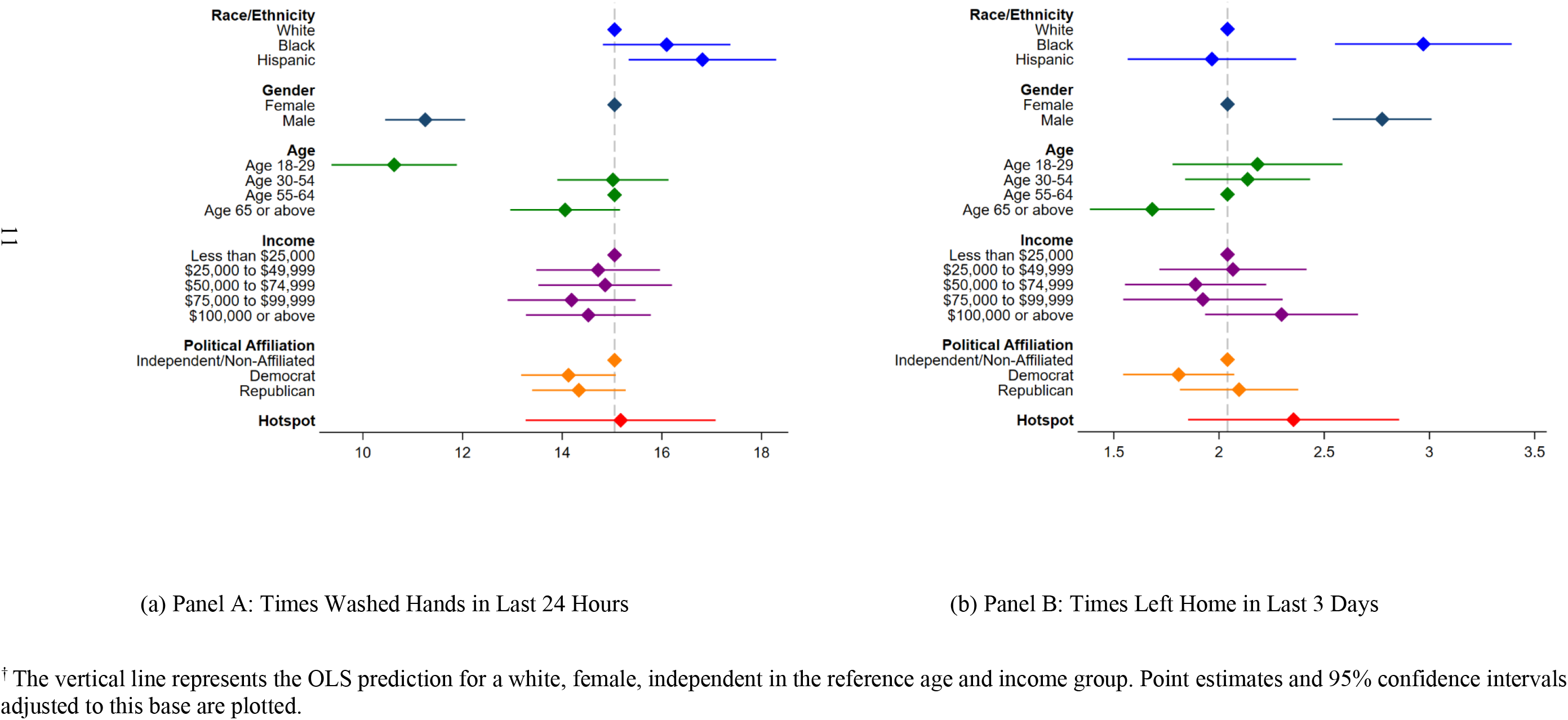
Hygiene and Social Distancing Behaviors^†^

## DISCUSSION

For society to control the spread of COVID-19, it is essential that public health officials know which groups are most heavily affected by the disease and that all individuals have access to trusted guidance on how to protect themselves and their communities from infection. Given the increased vulnerability to COVID-19 among low income households and racial/ethnic minorities, it is important to determine how accurate knowledge and public health behavior regarding COVID-19 is among these groups. Further, there is concern that a partisan divide in COVID-19 associated news coverage has weakened the effectiveness of public health messaging overall.

The paper uses national survey data conducted in the first few weeks of the COVID-19 pandemic in the United States to discern reported exposure, knowledge, and behaviors regarding its spread. The study reaches three conclusions. First, similar to what has been documented in the media, African-Americans are more likely to report COVID-19 exposure or know someone who has been infected. Reported COVID-19 exposure is also higher for men and younger age groups.

Second, although knowledge about the novel virus and how to reduce its spread is relatively high, there are important differences across groups that map to behaviors. In the population as a whole, over 80% of people know that they can contract COVID-19 from a contaminated surface, and even higher shares know the three cardinal signs of having COVID-19. Such high levels of knowledge for a novel disease testify to the role of public health officials and the news media in explaining the basics of disease spread. Yet African Americans, men and younger people, the very same groups which report higher COVID-19 exposure, have less accurate knowledge than white Americans, women and older individuals. The gaps are consistently largest for African-American respondents who are less likely to know about fomite transmission, asymptomatic transmission, respiratory droplet transmission or name three common symptoms of COVID-19.

Third, there were important differences in reported behavior. African-Americans, men and younger aged individuals are more likely to leave their homes. Some of the difference in behavior may be related to social circumstance. For example, African-Americans less likely to be able to telecommute and more likely to work in the service sector and use public transportation than other racial/ethnic groups (23% among African-Americans vs. 15% among Hispanics and 7% among whites).^14^ Strikingly, knowledge and behaviors are closely related; groups where behaviors put people more at risk for disease are also groups where knowledge of appropriate behaviors are weakest. The results suggest that greater efforts to communicate risk and safe practices to racial/ethnic minorities and younger people may be particularly crucial moving forward.

### Limitations

The study has several limitations. First, knowledge and behaviors associated with COVID-19 are changing rapidly, and it is possible that the findings here will be less applicable over time. Second, the survey was conducted by internet, which may bias the results towards those with access to Wi-Fi. Third, the results on reported COVID-19 infection did not specify whether the person had been tested for the disease.

### Conclusion

More effort will be needed to address the knowledge and behavior gaps between whites and African-Americans, between men and women, and between older and younger populations.

## Data Availability

The data are not currently publicly available.

## Acknowledgement

We are grateful to Joyce Kim, Sarah Eichmeyer, Luca Bragheri and Archie Hall for research support and Dynata for assistance with the survey.

## References

1 Garg, S, Kim L, Whitaker M, et al. Hospitalization rates and characteristics of patients hospitalized with laboratory-confirmed coronavirus disease 2019 — COVID-NET, 14 States, March 1–30, 2020. Morbidity and Mortality Weekly Report 2020; 69: 458-464. (Accessed April, 2020, at https://www.cdc.gov/mmwr/volumes/69/wr/pdfs/mm6915e3-H.pdf)

2 Chinni D. Coronavirus risk for African Americans tied to more than race. NBC Universal News Group 2020. (Accessed April 12, 2020, at https://www.nbcnews.com/politics/meet-the-press/coronavirus-risk-african-americans-tied-more-race-n1182146)

3 Thebault R, Ba Tran A, Williams V. The Coronavirus is infecting and killing black Americans at an alarmingly high rate”. The Washington Post. WP Company 2020. (Accessed April 7, 2020, at https://www.washingtonpost.com/nation/2020/04/07/coronavirus-is-infecting-killing-black-americans-an-alarmingly-high-rate-post-analysis-shows/?arc404=true)

4 Stafford K, Hoyer M, Morrison A. Outcry over racial data grows as virus slams black Americans. AP NEWS. Associated Press 2020. (Accessed April 8, 2020, at https://apnews.com/71d952faad4a2a5d14441534f7230c7c)

5 Lardieri A. Hispanics and blacks are hardest hit by COVID-19 in New York City. US News 2020. (Accessed April 8, 2020, at https://www.usnews.com/news/national-news/articles/2020-04-08/coronavirus-disproportionally-kills-hispanics-and-blacks-in-new-york-city)

6 Gupta S. Why African-Americans may be especially vulnerable to COVID-19. ScienceNews 2020. (Accessed April, 2020, at https://www.sciencenews.org/article/coronavirus-why-african-americans-vulnerable-covid-19-health-race)

7 Maqbool A. Coronavirus: Why has the virus hit African Americans so hard? BBC News 2020 (Accessed April, 2020, at https://www.bbc.com/news/world-us-canada-52245690)

8 Wong MD, Shapiro MF, Boscardin WJ, Ettner SL. Contribution of major diseases to disparities in mortality. New England Journal of Medicine 2002; 347(20):1585–92.

9 Boulware LE, Cooper LA, Ratner LE, LaVeist TA, Powe NR. Race and trust in the health care system. Public health reports 2016.

10 Alsan M, Wanamaker M. Tuskegee and the health of black men. The Quarterly Journal of Economics 2018; 133(1):407–55.

11 Green TV, Tyson A. 5 Facts about partisan reactions to COVID-19 in the U.S. Pew Research Center 2020. (Accessed April, 2020, at https://www.pewresearch.org/fact-tank/2020/04/02/5-facts-about-partisan-reactions-to-covid-19-in-the-u-s/)

12 Allcott H, Boxell L, Conway J, Gentzkow M, Thaler M, Yang DY. Polarization and Public Health: Partisan Differences in Social Distancing during the Coronavirus Pandemic. NBER Working Paper 2020; w26946.

13 Thompson SA, Serkez Y, Kelly L. How has your state reacted to social distancing? The New York Times 2020. (Accessed April, 2020, at https://www.nytimes.com/interactive/2020/03/23/opinion/coronavirus-economy-recession.html)

14 Anderson M. Who relies on public transit in the U.S. Pew Research Center 2016. (Accessed April, 2020, at https://www.pewresearch.org/fact-tank/2016/04/07/who-relies-on-public-transit-in-the-u-s/)

